# Treatment Attrition, Concomitant Pharmaceutical Use, Temporal Claims-Based Utilization, and Real-World Characteristics Among Esketamine New Initiators in the United States, 2019-2022

**DOI:** 10.1101/2024.04.23.24306221

**Authors:** Brandon G. Truax

## Abstract

**Aims:** To investigate real-world treatment attrition, concomitant use of pharmaceuticals with known drug-interactions, total cost of care, claims-based outcomes, and health disparities for individuals initiated on esketamine therapy in the United States.

**Methods:** Individuals aged 18-64 with >=1 adjudicated claim for esketamine intranasal spray from 2019 to 2022 in the Optum Labs Data Warehouse (OLDW) were included. Continuous health plan enrollment was required for 6 months prior and 6 months post-esketamine start date. Medical claims, pharmacy claims data, and socioeconomic data were descriptively analyzed to investigate characteristics of the real-world esketamine cohort.

**Results:** There were 833 individuals in the esketamine analysis cohort. 33% had <8 treatment sessions and did not finish the first stage of treatment (induction phase). Use of pharmaceuticals with drug interactions was high-60.2% had at least one prescription fill for a drug with a known esketamine interaction in the 90-day window after esketamine initiation. Total costs of care went from $2,905 per patient per month (PPPM) in the baseline period to $5,734 PPPM in the follow-up period. Emergency department utilization with mental-health related diagnoses reduced by 42.5% in the follow-up period. PPPM utilization for office visits, excluding all claims on esketamine treatment days, went up 45% in the follow-up period. The esketamine cohort was pharmaceutically complex, and many of them had fractured care: 20% had prescription fills for >=11 drugs in the follow-up period. 18.6% percent of the cohort had >=7 drug-prescribing providers, and over 35% of the cohort had encounters with more than 10 healthcare practitioners in the follow-up period. The esketamine cohort had 26.6% more individuals in the highest socioeconomic status quintile (least socially deprived), compared to individuals with treatment-resistant depression (TRD), not initiated on esketamine.

**Limitations:** This was a retrospective cohort descriptive analysis with small sample sizes. Additional statistical analysis was not performed.

**Conclusions:** The esketamine cohort was characterized by considerable complexity from both a polypharmacy and lifestyle perspective. These contextual factors likely had significant bearing on adoption, access, and eventual claims-based outcomes. Understanding the interplay of these factors with the treatment dynamics of consciousness-altering compounds will be important to obtain the best real-world results from therapeutic classes with psychedelic compounds.

## Introduction

Esketamine was approved by the United States (U.S.) Food and Drug Administration (FDA) in March 2019 as a therapeutic agent for treatment-resistant depression. Treatment guidelines for TRD recommend esketamine as an option for patients who have not responded to several pharmacologic trials.^1^ The terms of FDA’s Risk Evaluation and Mitigation Strategy (REMS) require esketamine be administered in an office setting under supervision of a healthcare professional. To date, real-world assessments of esketamine use have been limited to single-center cohorts and clinical sub-populations.^2,3^ Prior analyses have not investigated key factors that impact adoption, outcomes, and value such as treatment attrition, health disparities, and total healthcare resource utilization.

FDA approval of esketamine marks a paradigm shift that views compounds with psychedelic and consciousness-altering properties as potential therapies for mental health disorders. As of November 2023, there were more than 270 psychedelic drugs in various stages of development.^4^ Methylenedioxymethamphetamine (MDMA), a non-classical psychedelic, and psilocybin, a classical psychedelic, have gained ‘breakthrough’ status by FDA, putting the drugs on a regulatory fast-track to approval. While some of these compounds have been used for millennia by indigenous peoples and various cultures around the world, medicalization in the formal health system is just now coming to the fore.^5^

To better characterize outcomes and utilization for these nascent treatments, this analysis describes the real-world characteristics of individuals treated with esketamine. It explores how well they stick to their esketamine treatment plans, use of other medications before and after starting esketamine, and how often they use healthcare services. This study focuses on adults in the U.S. who have commercial health insurance.

## Methods

### Data Source

Administrative claims data from the OptumLabs^®^ Data Warehouse (OLDW) were used for analysis. The OLDW is a longitudinal, real-world data asset with de-identified administrative claims and electronic health record (EHR) data.^6^ This retrospective cohort study was determined to be exempt from institutional review board evaluation by the UnitedHealth Group Office of Human Research Affairs because it was a retrospective analysis of de-identified data. The analysis followed the STROBE reporting guidelines for cohort studies.

### Analysis Design

This retrospective cohort descriptive study encompasses data from January 2018 – December 2022. The index date window began with FDA approval of esketamine in March 2019. Enrollment and healthcare utilization data were pulled six months prior to the index date (baseline period) and six months post the index date (follow-up period).

### Analysis Cohort

This study looks at people who have health insurance and have used esketamine nasal spray at least once during a specific time. Esketamine is mainly prescribed for TRD so it’s likely that most people using it have TRD. This assumption is based on the fact that health insurance plans usually require approval before covering esketamine due to its cost and the rules around its use. Also, there’s a common practice of using a cheaper, generic version of ketamine off-label for similar purposes.

### Sample Selection

The data analysis included individuals with at least one adjudicated medical or pharmacy claim with esketamine. Esketamine claims processed through the medical benefit were identified using Current Procedural Terminology® (CPT) codes – S0013, G2082, and G2083. Pharmacy claims for esketamine were identified using National Drug Codes (NDCs) – 50458002800, 50458002802, and 50458002803.

#### Outcomes measures and definitions

To investigate utilization before and after esketamine initiation, individuals were included in the analysis if they had 12 months of continuous enrollment with medical and pharmacy coverage; 6 months before esketamine initiation (baseline period), and 6 months after esketamine initiation (follow-up period). This design permitted all individuals in the cohort the opportunity to complete the one-month esketamine induction phase (treatment sessions 1-8) and enter the maintenance phase of esketamine therapy (treatment sessions 8-12) in month two and beyond. Treatment attrition and completion was measured by the number of esketamine treatment dates.

Utilization of pharmaceuticals with known drug interactions (amphetamines, benzodiazepines, monoamine oxidase inhibitors, and opiate agonists), that may inhibit treatment response or increase adverse events, were defined the by prescribing information on FDA’s labeling insert for esketamine. American Hospital Formulary Service (AHFS) mappings were used for grouping generic drugs into their respective therapeutic classes. Utilization was defined by >=1 fill date for pharmacy claims from one or more of the drug classes. Analyses focusing on utilization of medications with drug interactions centered on the 90-day window prior to esketamine initiation or the 90-day window after esketamine initiation. A shorter time window was chosen relative to longer baseline and follow-up periods because it is more likely to accurately capture the simultaneous use of other medications alongside esketamine treatments.

Total costs of care and per patient per month (PPPM) costs were calculated by summing out-of-pocket and health plan paid amounts. Healthcare costs were inflation-adjusted utilizing the medical component of the US Consumer Price Index and are presented in 2022 dollars. We broke cost into six categories: emergency department, inpatient, office visits, outpatient, pharmacy, and other medical claims cost.

Emergency department visits were defined as revenue codes 0450-0459 or 0981, CPT codes 99281, 99282, 99283, 99284, 99285, 99288; with mental-health related visits including claims with International Statistical Classification of Diseases and Related Health Problems® (ICD-10) diagnoses F01 through F99, and all other emergency department visits considered non-mental health related.

Concomitant drug use, and pharmaceutical complexity (polypharmacy), was described as the number of distinct drugs with fill dates. This was defined by the number of different drugs, at the generic ingredient granularity, with prescription fill dates in the specified analysis windows.

The number of different drug-prescribing providers and total number of different providers with visits, were calculated per member, in the baseline and follow-up periods.

A sub-analysis aimed to investigate health disparities and used social deprivation index (SDI) data from the Robert Graham Center. The SDI draws on results from the American Community Survey (ACS) as an approximate measure for social determinants of health. The esketamine treatment cohort was compared to a major depressive disorder (MDD) with TRD cohort without an esketamine claim (see Appendix A). MDD TRD was defined as having >=1 primary, secondary, or tertiary diagnosis with ICD-10 CM F32.X or F33.X and having filled >= three antidepressants in a 12-month period.

#### Statistical Analysis

All analyses were descriptive in nature-using means, absolute counts, relative percentages, and frequency distributions. Rigorous statistical analysis and modeling was not undertaken.

## Results

### Cohort characteristics

The analysis included 833 individuals, aged 18-65, with >= 1 medical or pharmacy claim for esketamine (Table 1). 489 [58.7%] were female and 344 [41.3%] were male; mean age was 41.6 years.

**Table 1.**
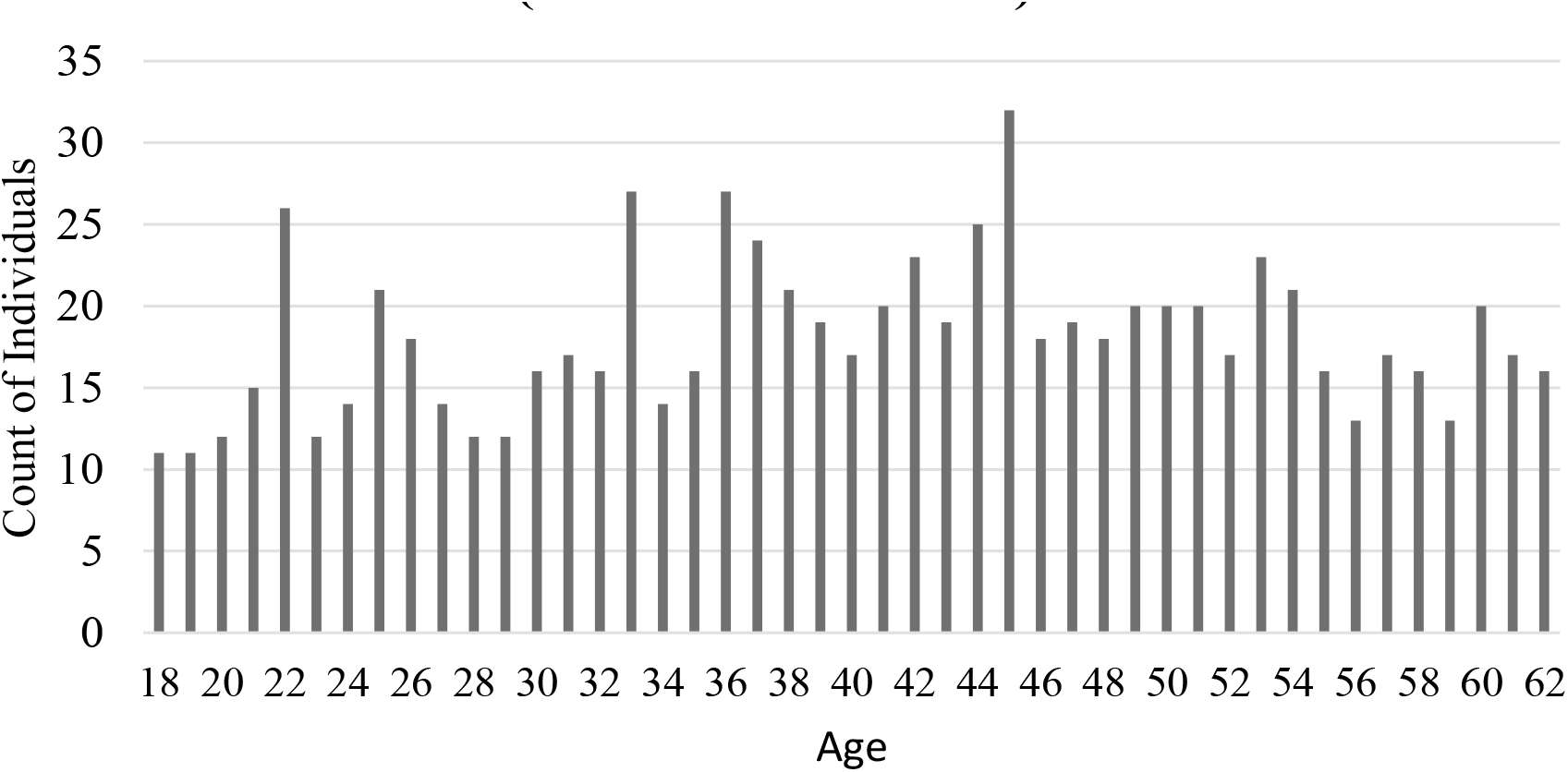
Age Distribution for Esketamine Cohort (% Female: 58.7%)

### Esketamine treatment attrition

Esketamine protocol completion was measured by the number of esketamine treatment session dates (Table 2); 69 individuals [8.3%] had 1 treatment session dates, 106 [12.7%] 2-4, 100 [12.0%] 5-7, and 89 [10.7%] 8-10, 78 [9.4%] 11-13, 91 [10.9%] 14-16, 300 [36%] 17+.

**Table 2.**
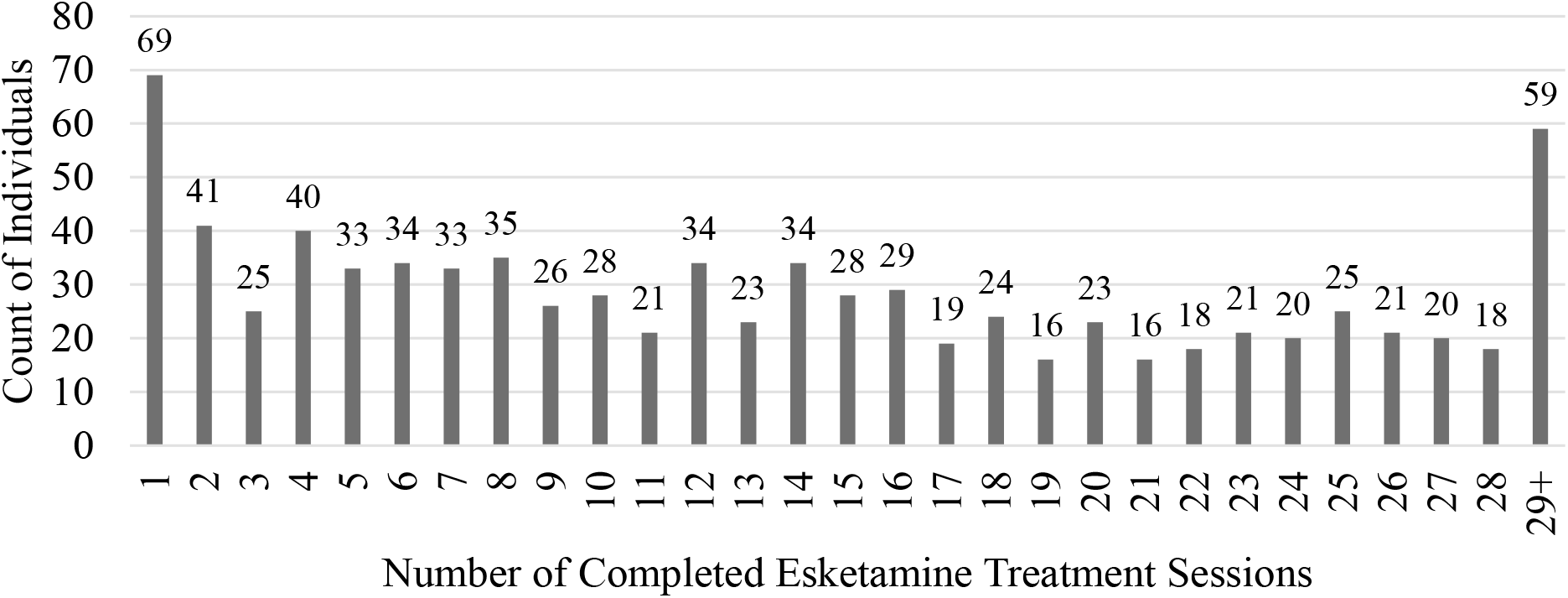
Count of individuals by number of completed esketamine treatment sessions over first six months.

### Utilization of pharmaceuticals with potential drug interactions

In the 90 days prior to esketamine therapy initiation, 526 [63.1%] filled at least one medication that has known drug interactions with esketamine (benzodiazepines, amphetamines, opiate agonists, or monoamine oxidase inhibitors) (Table 3). In the 90 days post-esketamine initiation, 502 [60.2%] filled at least one medication that has known drug interactions with esketamine.

**Table 3.**
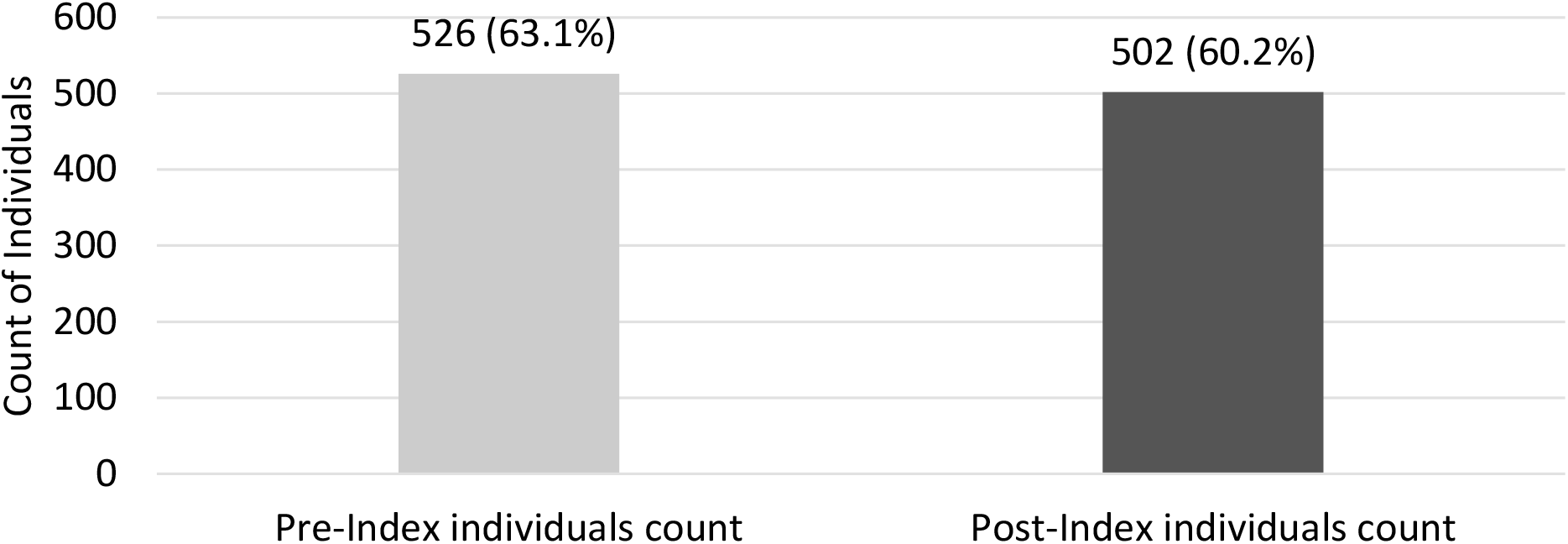
Utilization of Pharmaceuticals with Possible Drug Interactions In 90 day Period (pre and post esketamine initiation)

Of the individuals in the esketamine cohort, 201 individuals had a prescription fill in the amphetamines AHFS therapeutic class [24.1%], 172 benzodiazepines (anticonvulsants) [20.6%], 250 benzodiazepines (anxiolytic/sedative) [30.0%], 104 opiate agonists [12.5%] (Table 4). In the 90 days post-esketamine initiation, 198 individuals had a prescription fill in the amphetamines AHFS therapeutic class [23.8%], 159 benzodiazepines (anticonvulsants) [19.1%], 227 benzodiazepines (anxiolytic/sedative) [27.3%], 114 opiate agonists [13.7%].

**Table 4.**
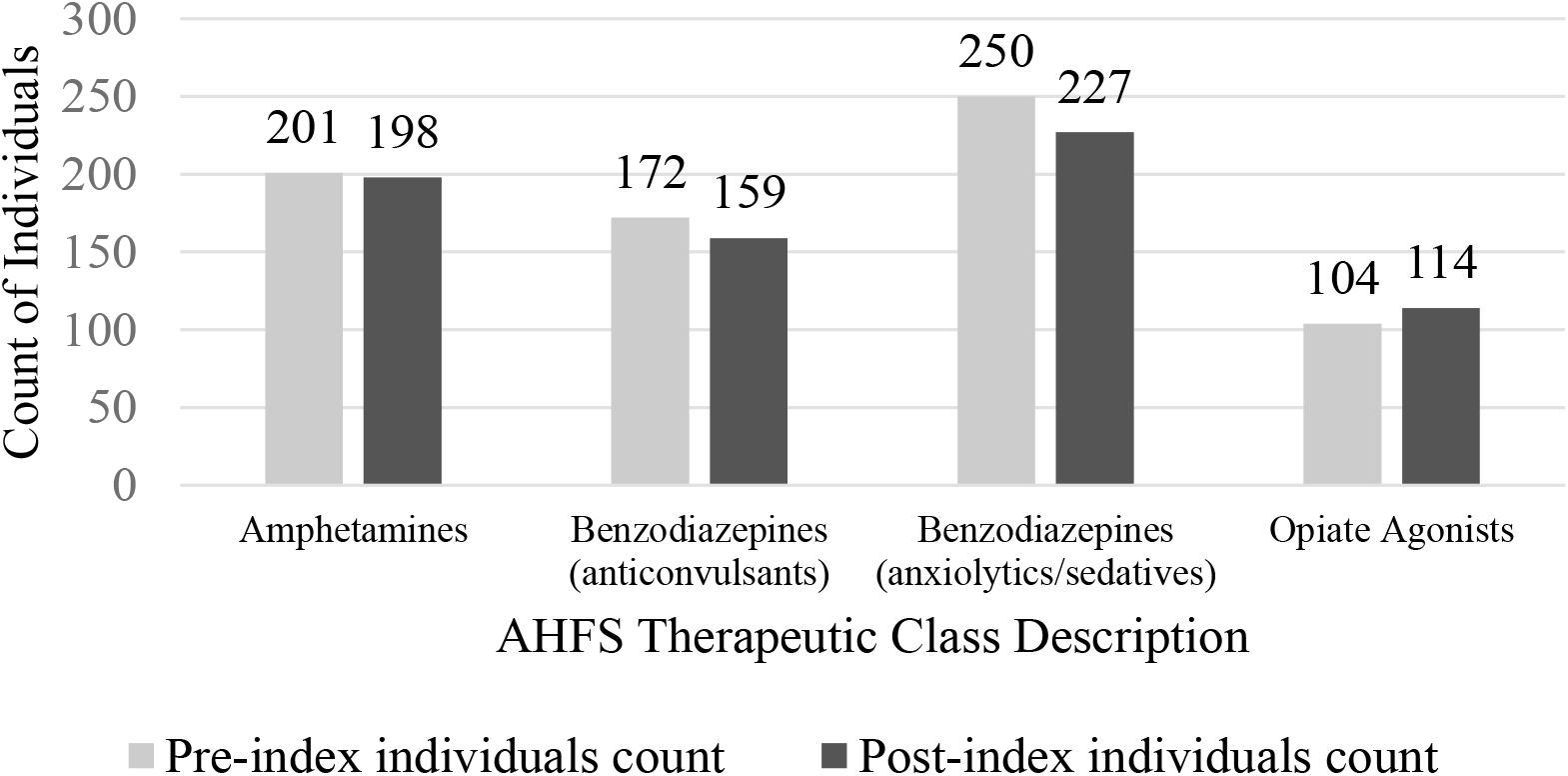
Utilization of Pharmaceuticals with Possible Drug Interactions in 90 Days Pre and Post Esketamine Start, by AHFS Class.

### Total cost of care and Healthcare resource utilization

In examining healthcare resource utilization (Table 5), total cost of care, partitioned by medical and pharmacy claims categories, was analyzed for the baseline and follow-up periods. Medical claims PPPM cost was $2,142 in the baseline period and $2,790 in the follow-up period. PPPM pharmacy claims cost was $763 in the baseline period and $2,944 in the follow-up period.

**Table 5.**
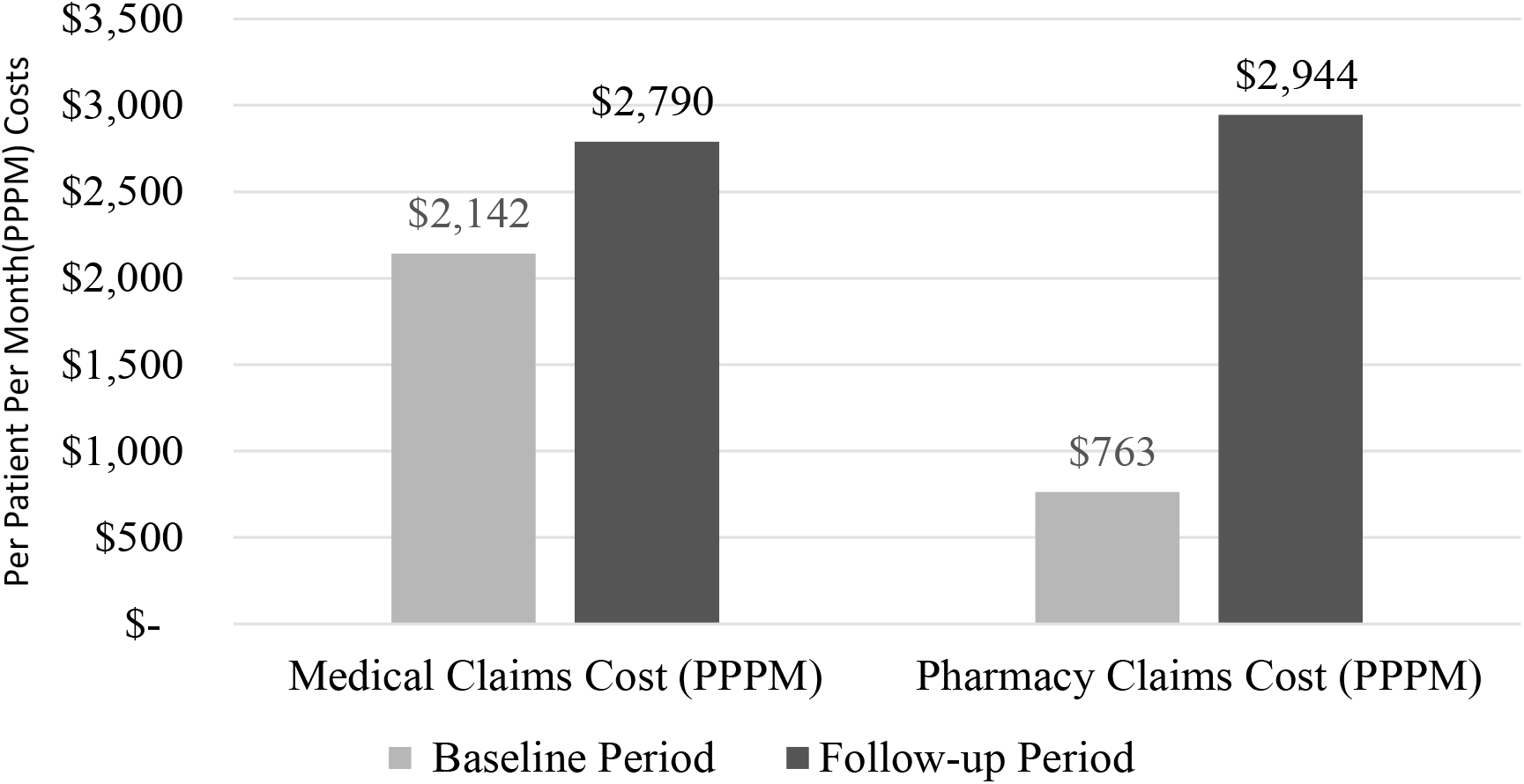
Total Cost of Care (medical and pharmacy claims) for Esketamine Cohort, Pre and Post-Index Date.

Excluding all medical and pharmacy claims costs on esketamine treatment dates in the follow-up period (Table 6), PPPM costs by service category were analyzed (Table 6). In the baseline period, PPPM costs were $588, outpatient $544, emergency department $209, office visits $579, other-medical $222, and pharmacy claims costs $763. In the follow-up period, inpatient PPPM costs were $688, outpatient $463, emergency department $200, office visits $840, other-medical $164, and pharmacy claims $723. On aggregate, excluding all esketamine treatment day-related utilization, PPPM costs (medical and pharmacy) were $2,905 in the baseline period, and $3,078 in the follow-up period.

**Table 6.**
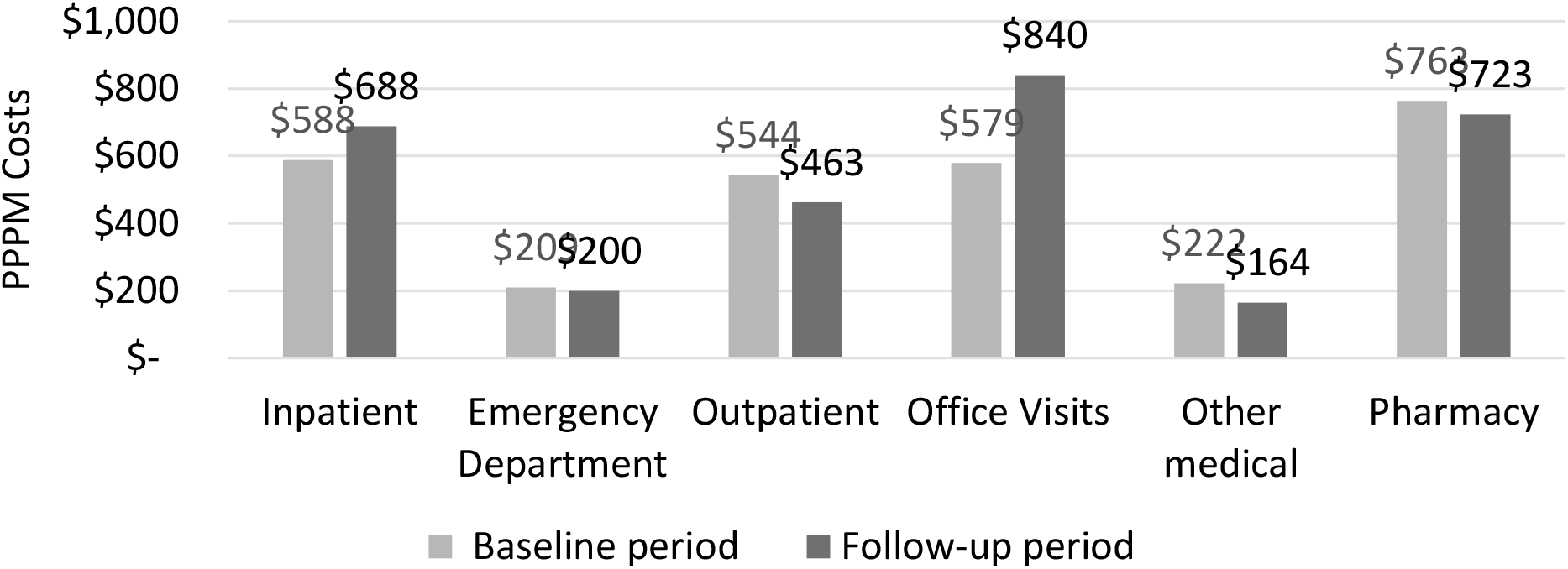
Healthcare Resource Utilization by Service Category; Pre-index and Post-index.

### Emergency Department Utilization

Emergency department (ED) utilization was measured by ED visits per 1,000 individuals (Table 7). In the baseline period, there were 96 mental health-related ED visits/1,000 individuals and 337 non-mental health-related visits/1,000 individuals. In the follow-up period, there were 55 mental health-related ED visits/1,000 members and 331 non-mental health-related visits.

**Table 7.**
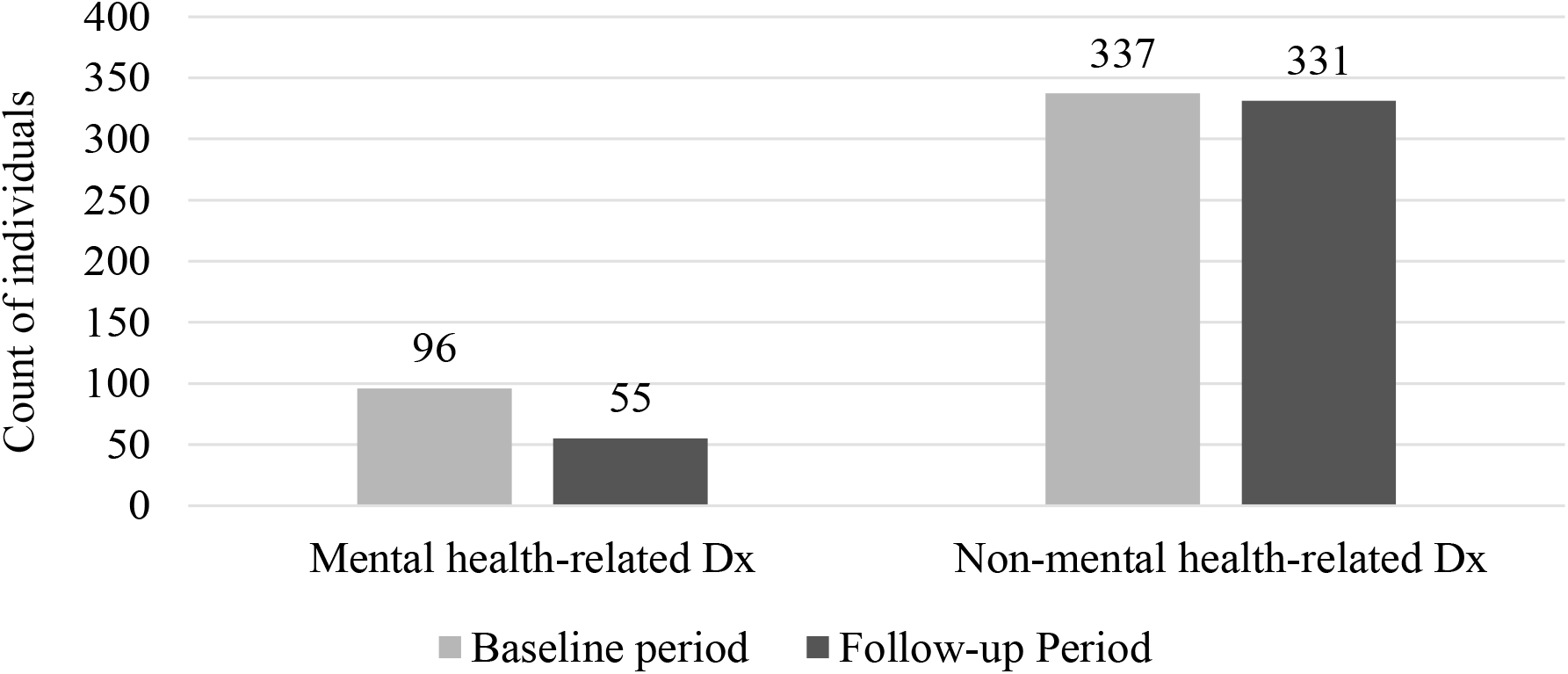
Emergency Department Service Dates Per 1,000 Individuals.

### Concomitant drug use and pharmaceutical complexity

In the baseline period, 328 individuals [39.4%] had prescription fills for 0-5 drugs, 326 [39.1%] 6-10 drugs, 179 [21.5%] 11+ drugs (Table 8). For the follow-up period, 357 individuals [42.9%] had prescription fills for 0-5 drugs, 304 [36.5%] 6-10 drugs, 172 [20.6%] 11+ drugs.

**Table 8.**
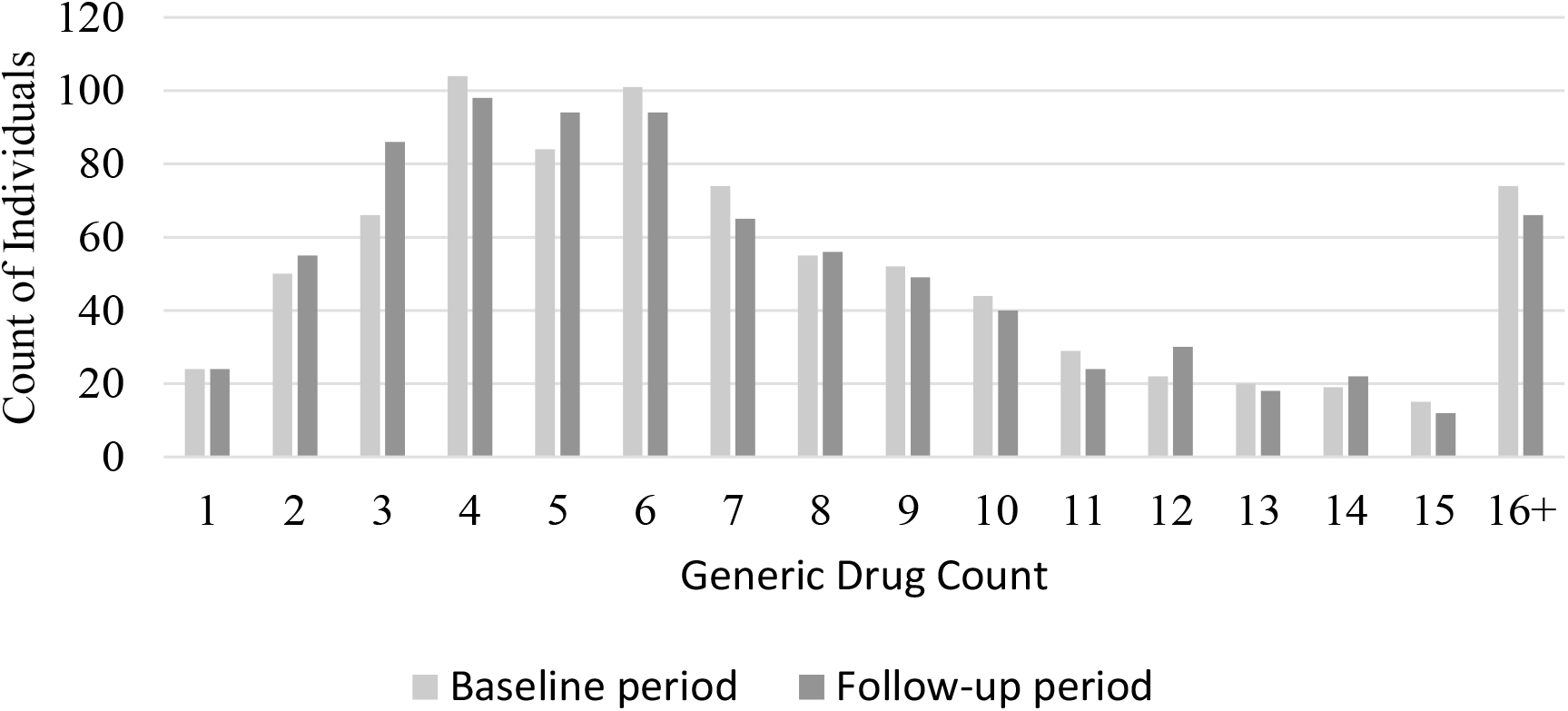
Number of Different Generic Drugs with Prescription Fill Dates.

### Number of drug-prescribing providers

In the baseline period, 377 individuals [45.2%] had prescription fills prescribed by 0-3 distinct healthcare providers, 302 [36.3%] 4-6 providers, and 154 [18.5%] 7+ providers(Table 9). For the follow-up period, 339 individuals [40.7%] had prescription fills by 0-3 providers, 339 [40.7%] 4-6 providers, and 155 [18.6%] 7+ providers.

**Table 9.**
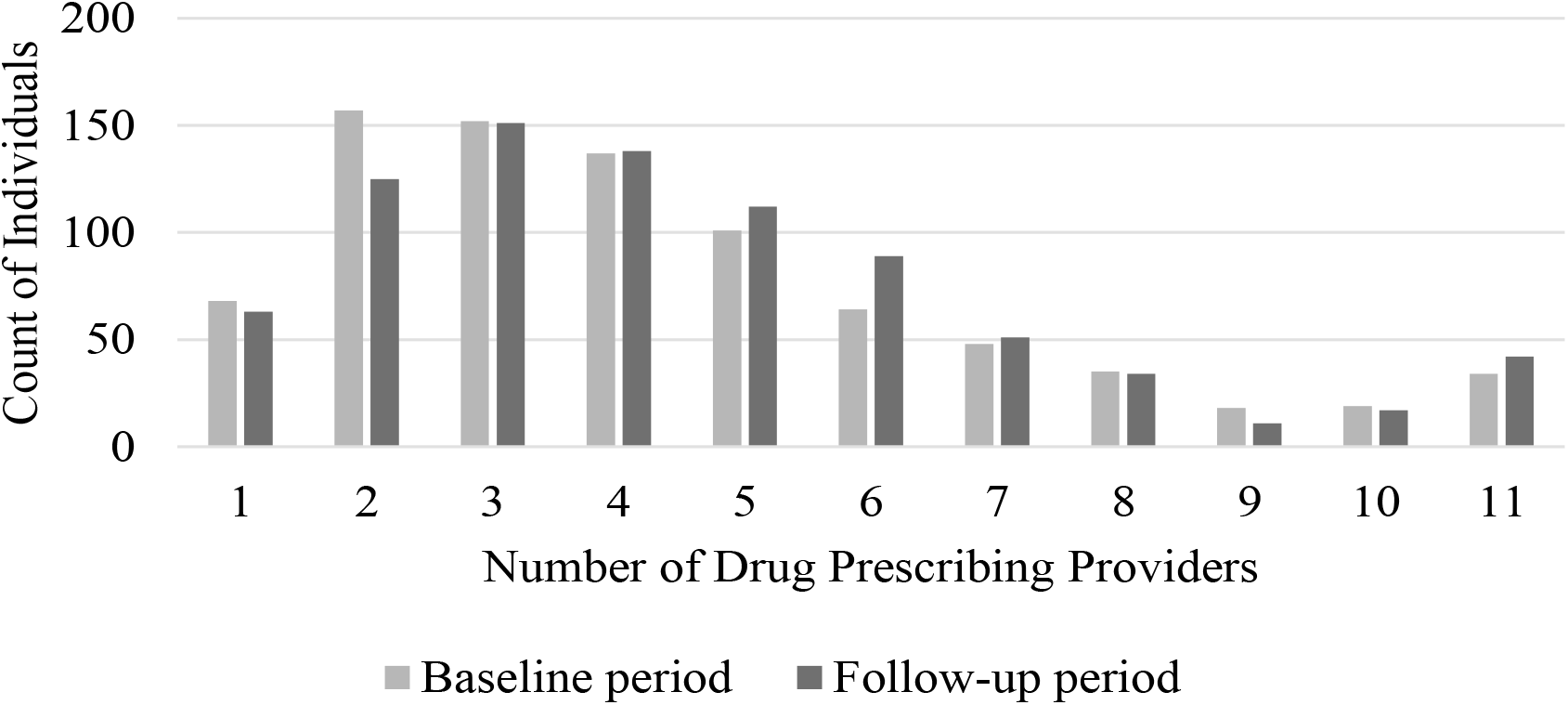
Count of Distinct Drug-Prescribing Healthcare Providers.

### Number of different providers with encounters

In the baseline period, 208 individuals [25.0%] had medical claims with 0-4 healthcare providers, 303 [36.4%] 5-9 providers, and 322 [38.7%] 10+ providers (Table 10). In the follow-up period, 199 individuals [23.9%] had a medical claim with 0-4 providers, 342 [41.1%] 5-9 providers, and 292 [35.1%] 10+ providers.

**Table 10.**
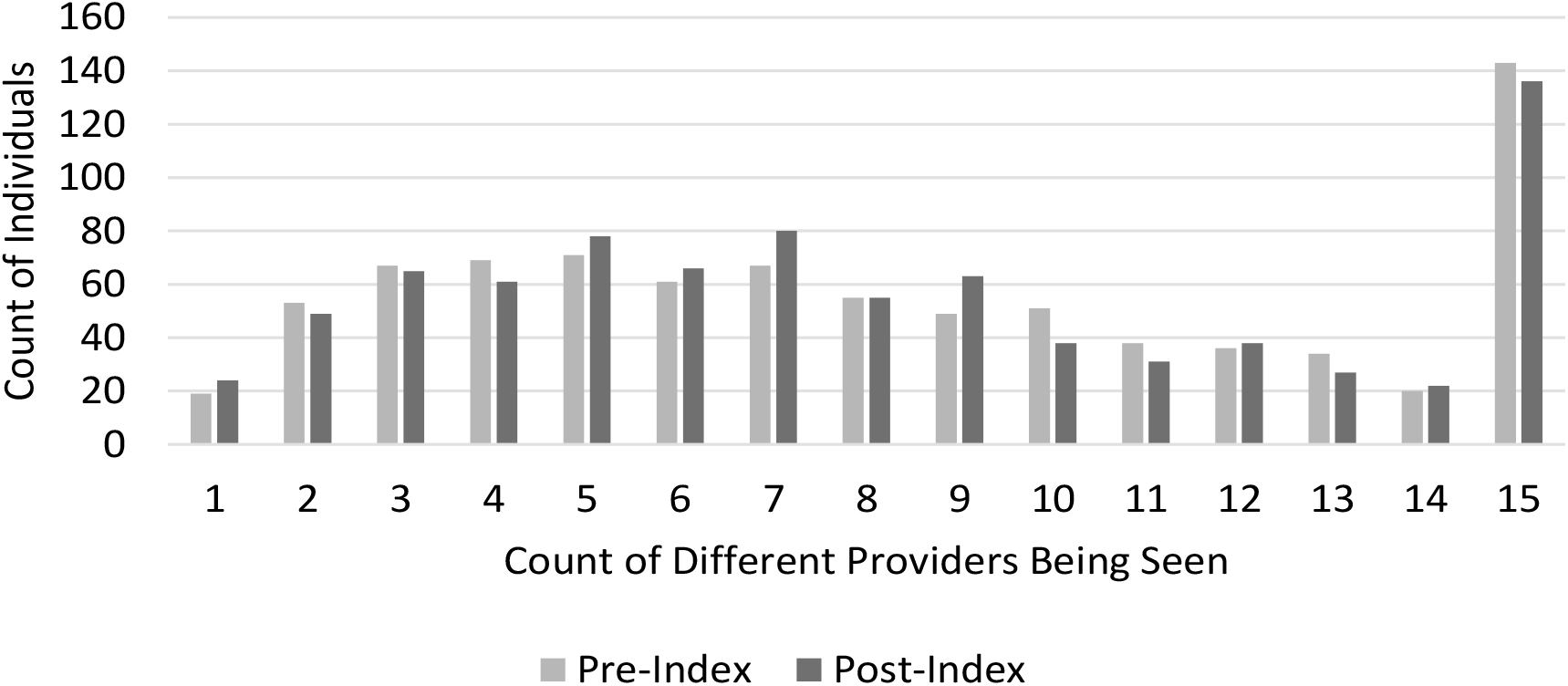
Count of Different Providers with Medical Encounters.

### Socioeconomic characteristics

Individuals in the MDD with TRD cohort (not initiated on esketamine) and the esketamine treatment cohort were grouped into SDI quintiles (Table 11). Quintile 1, SDI 1-20, represents individuals with the highest socioeconomic status (lease socially deprived); 30.8% of the TRD (no esketamine) cohort were in quintile 1, and 39.0% of the TRD (with esketamine) cohort were in quintile 1. Quintile 5, SDI 81-100, represents individuals with the most disadvantaged socio-economic status; 9.7% of the TRD (no esketamine cohort) were in quintile 5, and 8.8% of the TRD (with esketamine) were in quintile 5.

**Table 11.**
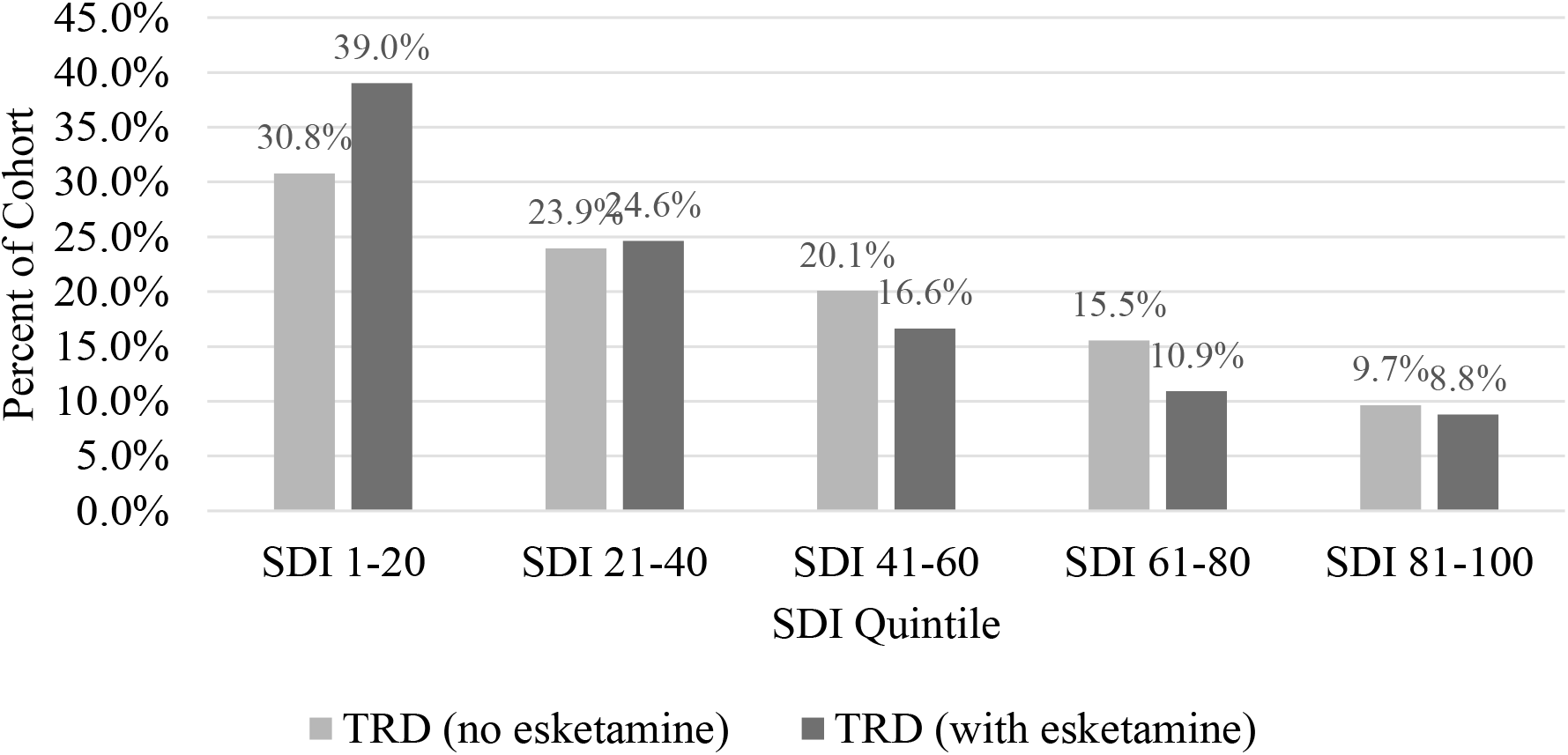
Percent of Individuals by Social Deprivation Index (SDI) For Each Analysis Cohort.

## Discussion

In the five years since esketamine’s initial FDA approval in 2019, a number of real-world dynamics have limited the reach and potential value of esketamine therapy.

First, 275 individuals (33%) ended esketamine treatment prior to finishing the induction phase and did not reach the esketamine maintenance phase (Table 2). This high attrition rate may represent multiple barriers to esketamine treatment adoption and continuation, including health plan coverage, out-of-pocket costs, logistics of participating in treatment (which requires >1 hour in a clinician’s office per session, and requires a ride home), safety and efficacy profiles, and sub-optimal treatment response or adverse events. This study focused on people with at least one completed claim and did not account for treatment access barriers to start esketamine treatment. Prior research has shown that 65 percent of initial esketamine claims may be rejected or abandoned due to plan coverage criteria, claims submission errors, or prior authorization restrictions.^2^ These barriers to treatment access may reduce the overall impact to the health system for esketamine and other therapies like it.

Second, using pharmaceuticals with known drug interactions with esketamine may put patients at risk for less effective treatment outcomes. In some cases, such as with benzodiazepines, drug interactions have been shown to decrease the effectiveness of esketamine therapy or elevate severity of adverse drug events.^7,8^ Due to the high degree of polypharmacy in the esketamine cohort (Table 8), there may be other drugs with interactions not investigated in clinical trials. More research is needed to understand esketamine treatment-response and effectiveness through stratifying individuals by heterogeneous polypharmacy sub-cohorts.

Total cost of care, including all medical and pharmacy claims, was $2,905 PPPM in the baseline period ($2,142 medical, $763 pharmacy) and $5,734 PPPM in the follow-up period ($2,790 medical, $2,944 pharmacy) (Table 5). This substantial increase in total claims costs in a follow-up window of 6 months, may inhibit adoption and reimbursement coverage for esketamine. Health payers and other risk-bearing entities have a limited time window to recoup treatment costs in their enrolled populations. ^9^ Time-to-breakeven and timely return on investment are often investigated with reimbursement and treatment coverage decisions by payers.

With regard to healthcare resource utilization by service category, results of esketamine treatment were mixed. A data sub-analysis excluded all claims costs on esketamine treatment dates in the follow-up period to see how HRU changed aside from any utilization on esketamine treatment days. PPPM costs in the office visits service category increased the most (+45%) from baseline period ($579) vs. follow-up period ($840) (Table 6). Future research could explore the factors in this spending growth.

In a claims-based utilization sub-analysis, mental health-related ED service dates decreased by 42.7% in the follow-up period, whereas non-mental health-related ED service dates decreased by only 1.8%. This data emphasizes the rapid-acting effect of esketamine on mental health-related emergencies, as cited in clinical trials and prior published research. These ED sub-analyses were not adjusted for condition severity or confounders.

Over 20% of individuals had prescription fills with more than 11 drugs after initiation of esketamine therapy. 18.6% of the esketamine cohort had 7 or more drug-prescribing providers in the follow-up period, and over 35% of individuals had medical encounters with 10 or more providers (Table 9, Table 10). This complexity of healthcare utilization imposes a significant load in patient management, makes unpredictable drug interactions more likely, and may result in sub-optimal care trajectories.^10^

People with TRD receiving esketamine therapy are of higher socioeconomic status compared to those with TRD that do not initiate esketamine treatment (Table 11). This may be due to myriad reasons: out-of-pocket costs, long treatment times with many visits, geographic constraints, or other logistical/systemic barriers. This potential for unequal treatment access is worrisome. If esketamine and other emerging psychedelic compounds become boutique therapies for socioeconomically advantaged people, their true benefits will be obscured in the usual jungle that confounds, complicates, and exacerbates health disparities.

As esketamine utilization grows, future research should focus on validating and reproducing real-world data insights across diverse patient populations: assessing safety and effectiveness outcomes, measuring cost-effectiveness, and defining predictors of treatment continuation and attrition. Characterizing additional features of real-world esketamine use will be necessary to optimize and re-design clinical care models to best suit patient needs.

### Limitations

This study has limitations, including those common to the use of administrative medical and pharmacy claims in retrospective cohort studies.^11^ Additionally, the study time period overlapped with COVID-19, which had a known influence on healthcare utilization and care-seeking behavior. The analysis cohort was comprised of people with commercial health insurance; further research is needed with people that have different health plan types such as Medicaid or Medicare. Due to insufficient sample size, this study was not designed to assess safety and efficacy outcomes or predict patient-level features associated with attrition. This analysis was exploratory and descriptive in nature, and rigorous statistical modeling was not performed.

## Conclusions

The pharmacology of esketamine, and its treatment protocol, are different than other consciousness-altering therapies and psychedelic compounds that are on the horizon to enter the health system. However, characteristics and real-world dynamics from the esketamine cohort may have implications for future psychedelic therapies as they are medicalized.

**Figure 1.**
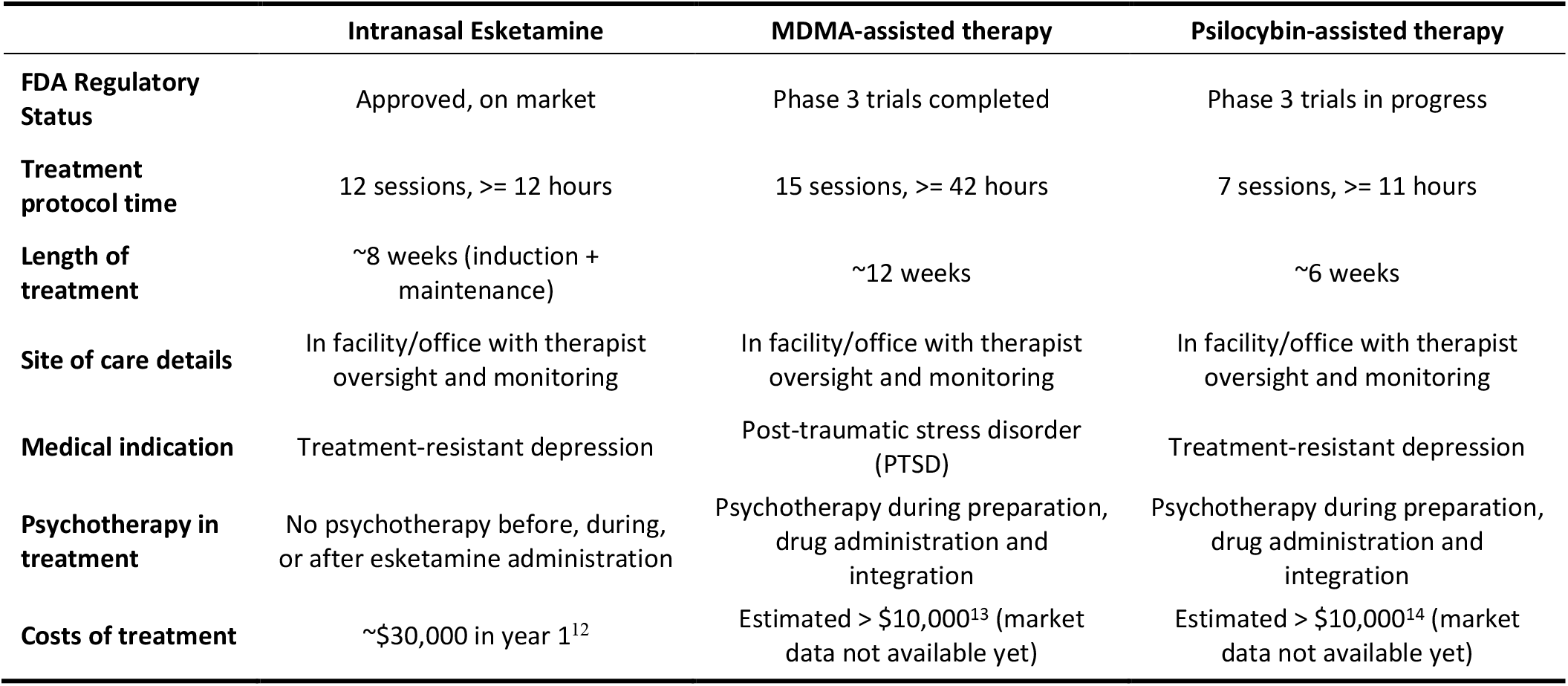
Comparison of treatment characteristics for consciousness-altering/psychedelic compounds.

Most all pharmaceuticals face a number of challenging real-world dynamics in the U.S. health system. However, certain access and system adoption headwinds for psychedelic compounds are amplified by the uniqueness of their current treatment models. Specifically, the high frequency of office visits requiring large amount of treatment time, significant short-term healthcare resource use and cost, and very medically complex patient cohorts are unique to this emerging therapeutic area.

To optimize benefit and reach, a number of factors will critically influence access to and outcomes of these nascent treatments:

1. Implementing strategies for transportation/logistical support and adaptable sites of care to promote treatment access and completion
2. Conducting regular comprehensive medication reviews to accurately identify patients with likely drug interactions and take into account concomitant drug profiles that may delay treatment response or cause adverse drug events.
3. Building equity and fairness into market access using outcomes-based agreements between drug manufacturers and risk-bearing entities with populations that are socioeconomically disadvantaged.
4. Running real-world pilots with payers and employers to optimally identify/stratify prospective patients— to prove clinical and financial value.
5. Proactively and frequently leveraging electronic health record transmission and timely provider-to-provider communication to minimize information asymmetry and potential for disjointed care.

Findings from this analysis highlight how complex underpinnings of the real-world health system can affect consciousness-altering treatment completion, clinical outcomes, and healthcare resource utilization. Optimizing health system implementation and effective scaling is necessary for potential individuals on esketamine. Real-world data research is essential to catalyze positive outcomes for patients that may initiate treatments with other consciousness-altering/psychedelic compounds in the future.

## Data Availability

The data is not publicly available due to data use agreement.

## Transparency

### Author Affiliation

University of St. Thomas, St. Paul, Minnesota

### Author contributions

BT conceived of the study, collection and assembly of data, and data analysis and interpretation.

## Appendix A

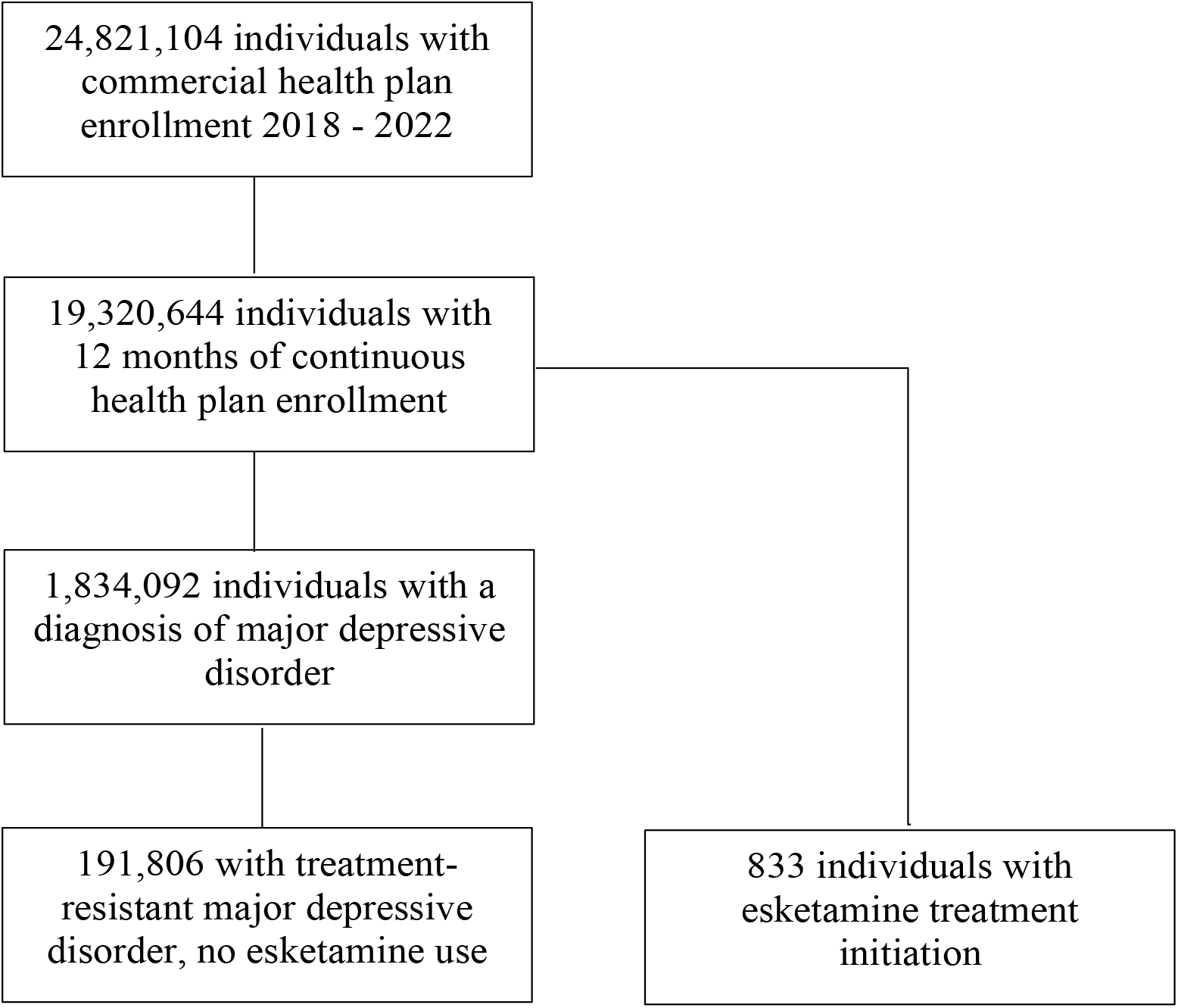

